# Pharmacometric evaluation of amodiaquine-sulfadoxine-pyrimethamine and dihydroartemisinin-piperaquine seasonal malaria chemoprevention in northern Uganda

**DOI:** 10.1101/2025.06.24.25330200

**Authors:** C. Bonnington, A. Nuwa, K. Theiss-Nyland, R. Kajubi, M. R. Kamya, J. Nankabirwa, C. Ebong, C. Rassi, J. Nabakooza, J. Opigo, D. Salandindi, M. Odongo, C. Sararat, J. A. Watson, K Suwannasin, S Proux, U Koesukwiwat, J Tarning, M Imwong, J. Tibenderana, FH Nosten, NJ White

## Abstract

**Background:** Seasonal malaria chemoprevention (SMC) using monthly amodiaquine-sulfadoxine-pyrimethamine (SPAQ) is being deployed East Africa, where antimalarial drug resistance levels are high. Dihydroartemisinin-piperaquine (DP) is a potential alternative.

**Methods:** A 28 day pharmacometric assessment of SMC was conducted in Karamoja, Northern Uganda. In two villages children received SPAQ and in one, either DP or no SMC. The primary outcome was malaria infection (all species) defined by capillary blood sample qPCR positivity on day 28, or new slide or rapid diagnostic test positivity after day 2.

**Results:** Baseline qPCR malaria parasitemia prevalence among 1250 enrolled children was 46% (575/1250); *P. falciparum* 85%, other malarias (mainly *P. ovale*) 25%. Breakthrough parasitemias occurred in 7% (33/496) of DP, 31% (158/504) of SPAQ, and 39% (98/250) of no drug recipients. Clinical malaria (all *P. falciparum*) developed in 17% of no drug (42/250; 1 severe), 8% of SPAQ (38/504; 2 severe) and 2% of DP (13/496) recipients. Adjusted protective efficacies against all malaria, all clinical malaria and all falciparum malaria were SPAQ; 56% (95%CI 35-70%), 60% (27-78%), and 46% (11-68%), and for DP; 84% (77-89%), 86% (74-92%) and 86% (75-92%) respectively. Some asymptomatic *P. falciparum* infections were not cleared by SPAQ. All 260 *P. falciparum* isolates genotyped were *Pfcrt* K76 (haplotype CVMNK, a marker of 4-aminoquinoline susceptibility) and most were quintuple *Pf dhfr/dhps* mutants (i.e. relatively SP resistant). The chemoprevention drug exposure-response relationship was strong for desethylamodiaquine, but weak for sulfadoxine.

**Conclusions:** SPAQ SMC had low clinical and parasitological chemopreventive efficacy in Northern Uganda whereas dihydroartemisinin-piperaquine was effective.

**Funding:** Bill & Melinda Gates Foundation; GiveWell; Wellcome. The Infectious Diseases Research Collaboration (IDRC) and Malaria consortium received funds from grant Investment IDs INV-043830 and INV-039889 - Uganda SMC and CPES Phase 2 from the Gates Foundation. The findings and conclusions contained within are those of the authors and do not necessarily reflect positions or policies of the Gates Foundation. NJW is a Principal Research Fellow funded by the Wellcome Trust (093956/Z/10/C). JAW is a Sir Henry Dale Fellow funded by the Wellcome Trust (223253/Z/21/Z).

**Clinical Trials registration:** NCT05323721

## Background

Seasonal malaria chemoprevention (SMC) is the administration of treatment doses of antimalarial drugs at monthly intervals to prevent malaria in areas of seasonal high malaria transmission. Originally SMC was deployed to children (aged between 3 and 59 months) for 3-5 months each year across the Sahel region of Africa, where falciparum malaria transmission is intense but highly seasonal [1–3]. In early 2022 the World Health Organization recommendations for both SMC and for perennial malaria chemoprevention (formerly IPTi) were broadened substantially and restrictions lifted on the number of monthly SMC cycles or the age of SMC recipients [4]. This new WHO guidance was not preceded by assessments of chemopreventive efficacy in East Africa, where falciparum malaria is more drug resistant and less seasonal. SMC usually comprises amodiaquine (for three days) plus single dose sulfadoxine-pyrimethamine (SPAQ) [1, 2, 5–10]. With reported elimination half-lives of approximately 3 (pyrimethamine), 7 (sulfadoxine) and 10 days (desethylamodiaquine), SPAQ is expected to provide effective antimalarial chemoprevention for one month [9, 11]. We assessed SPAQ SMC in Uganda and the potential alternative dihydroartemisinin-piperaquine (DP) by pharmacometric monitoring (PARM) [13].

## Methods

### Study location and design

This study was conducted in three large village areas in the Karamoja region, north-eastern Uganda - an area of intense malaria transmission with high levels of antimalarial drug resistance where SMC had not been deployed previously [14]. This was part of a larger clinical and implementation assessment reported elsewhere [12,15]. The implementation project was preceded by extensive training and community engagement. SPAQ was deployed in two villages and, in the third, children were randomized 2:1 to receive either DP or no drug.

### Screening and enrolment

Study supervisors used household lists compiled by the Village Health Teams to identify villages for the study. They then randomly selected 10 households per village with ≥ 1 eligible child which were visited to explain the study procedures. Only resident afebrile children, aged between 3-59 months with no symptoms of malaria in the past 48 hours, and whose caregivers were willing and able to comply with the study protocol throughout the study and provided written informed consent were included. Children were excluded if they had known allergies, had received recently drugs with antimalarial activity, were severely malnourished, HIV positive, or were included in other studies.

### Procedures

A baseline (day 0) weight, mid upper arm circumference (MUAC) and axillary temperature were recorded and capillary blood samples were collected. Observed single dose oral sulfadoxine-pyrimethamine (SP: infants 250/12.5mg; older children 500/25mg) was given, and amodiaquine (AQ: Infants: 75mg, older children 1-5 years: 150 mg) was given once daily for three days. In the third village DP (D-ARTEPP ® dispersible tablets, Fosun Pharma; 20mg dihydroartemisinin/160mg piperaquine) was given according to the WHO recommended weight-based dosing regimen. All illnesses within 28 days follow-up were assessed and details recorded. Treatment of uncomplicated malaria was oral artemether-lumefantrine (AL). Severe malaria treatment was parenteral artesunate or rectal artesunate followed by AL. Apart from the pharmacometric measurements [13], no additional interventions, monitoring or support was provided.

Malaria slides were prepared and dried blood spots (DBS) were stored on Whatman 31 ET Chromatography filter paper (Cat. No. 3031-915) on days 0, 7, 14, 21 and 28. After barcoding and dry storage they were airfreighted to the analytical laboratories in Thailand.

Detection and speciation of malaria was with qPCR [18,19] (approximate lower limit of quantitation [LLOQ] 500-1000 parasite genomes/mL). Blood slides from PCR identified recurrent infections were then examined by experienced microscopists. Punch blood spots were extracted and drug levels measured by validated LC MS/MS assays [16–18]; amodiaquine [AQ; LLOQ 1.87ng/mL], the biologically active metabolite desethylamodiaquine [DAQ; LLOQ 2.95ng/mL], pyrimethamine [PYM; LLOQ 4.42ng/mL], sulfadoxine [SDX; LLOQ 840ng/mL] and piperaquine [PIP; LLOQ 3ng/mL]).

The *primary outcome* was any malaria infection by day 28, either symptomatic malaria confirmed by rapid diagnostic test (RDT) or microscopy, or Day 28 blood spot PCR positivity (all species).

*Secondary outcomes* included clinical malaria by day 28, severe malaria, falciparum and non-falciparum malaria.

### Statistical analysis

The primary analysis was done according to treatment allocation (“intention to treat”) with a secondary (“per protocol”) analysis excluding children for whom drug measurement suggested protocol deviations. To adjust for differences between the villages in the force of infection, and thus immunity, baseline malaria positivity (slide and qPCR) and calendar date were used as proxies for spatiotemporal variations in force of infection, and age as a proxy for immunity in a multivariable Poisson regression model

For the pharmacometric assessment [13] the age-stratified relationship between day 7 or day 28 (depending on terminal elimination half-life; t_1/2_β) drug concentrations and qPCR positivity was characterised. The t_1/2_β was estimated from the log_10_ day 7 and 28 whole blood concentrations under a Bayesian hierarchical linear model with inter-individual variability in intercept and slope, accounting for left censoring for below LLOQ values. This systematically underestimates the piperaquine t_1/2_β which may not enter the terminal elimination phase before D7 [19].

*Ethical approval*: Mbale Regional Referral Hospital-REC (Ref: MRRH-2022-168) and the Uganda National Council for Science and Technology (UNCST) (Ref: HS2212ES). The study was sponsored by the Malaria Consortium.

## Results

### Overview

Between 11^th^ July and 2^nd^ September 2022 1250 children, median (range) age 36 (3 to 60) months, were studied; 504 were allocated to SPAQ; 496 to DP and 250 to no drug (sequential 2:1 allocation) (Figure 1). Median (IQR) z scores for weight and mid upper arm circumference (MUAC) were -0.62 (-1.43 to 0.23) and -2.47 (-3.13 to -1.29) reflecting a generally undernourished population (Table 1). Baseline malaria prevalences in the SPAQ villages (Karita: 129/253 [51%], Nakapiripirit: 162/251 [65%]) were higher than in the DP or no drug village (Lemusui: 284/746 [38%] (p<0.001). Otherwise, there were no significant differences between the three locations (Table 1). In general SMC was well tolerated. No serious adverse events were reported. Vomiting of SMC tablets was reported for 17 SPAQ and 2 DP recipients; risk ratio 8.4 (95%CI 1.9 to 36.0). Five children in the DP group had piperaquine in their D28 blood sample but not in the D7 sample. In the control group 27 children had piperaquine levels indicating that they had received DP incorrectly. These 32 children were considered unevaluable for the per-protocol analysis.

**Figure 1:**
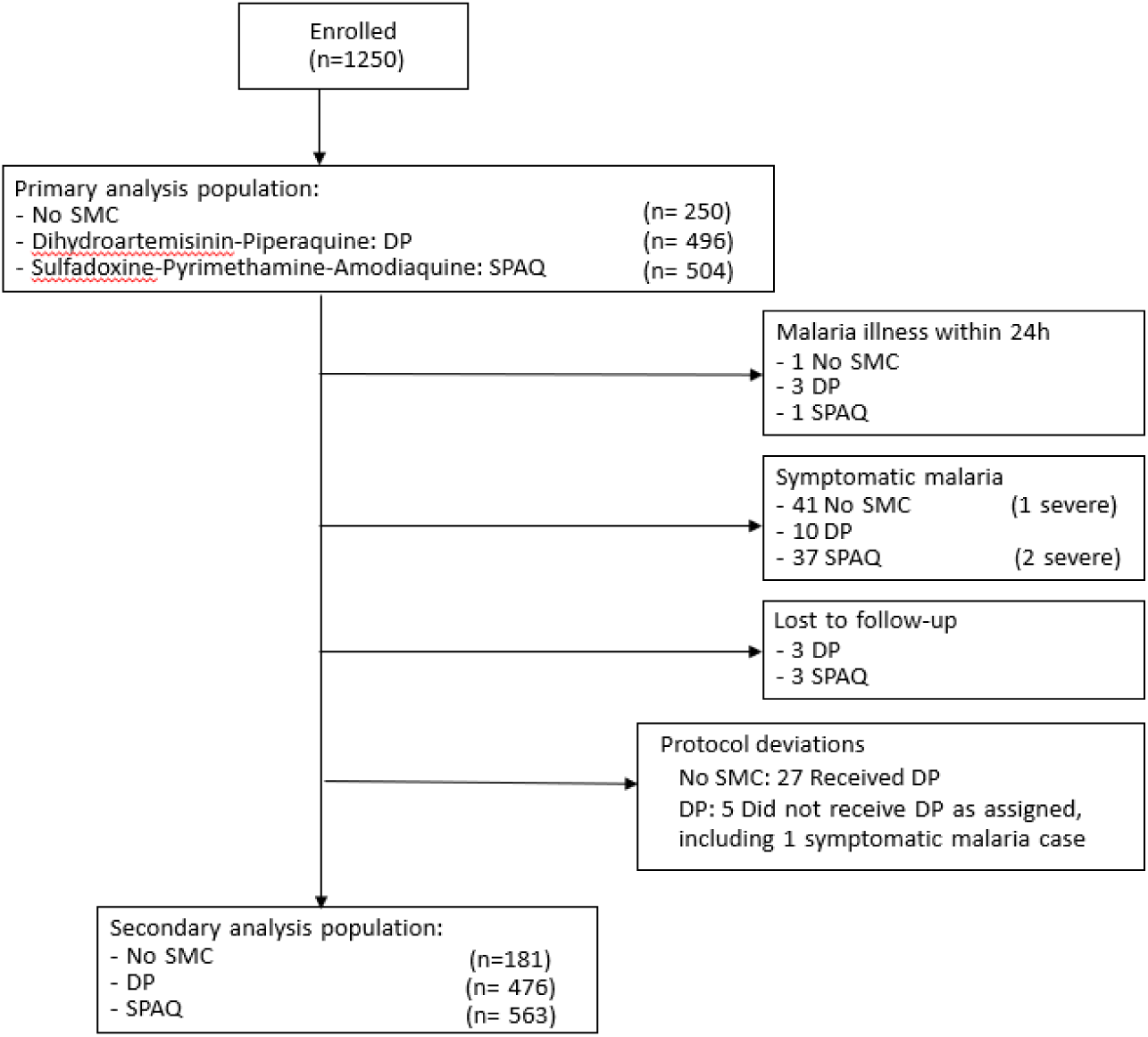
CONSORT diagram of participant flow in the trial Alt text for Figure 1: This is a CONDSORT diagram which shows the numbers of subjects enrolled, the numbers eligible for the primary analysis, their treatment allocations, and the numbers in the primary and secondary analyses.

**Table 1:**
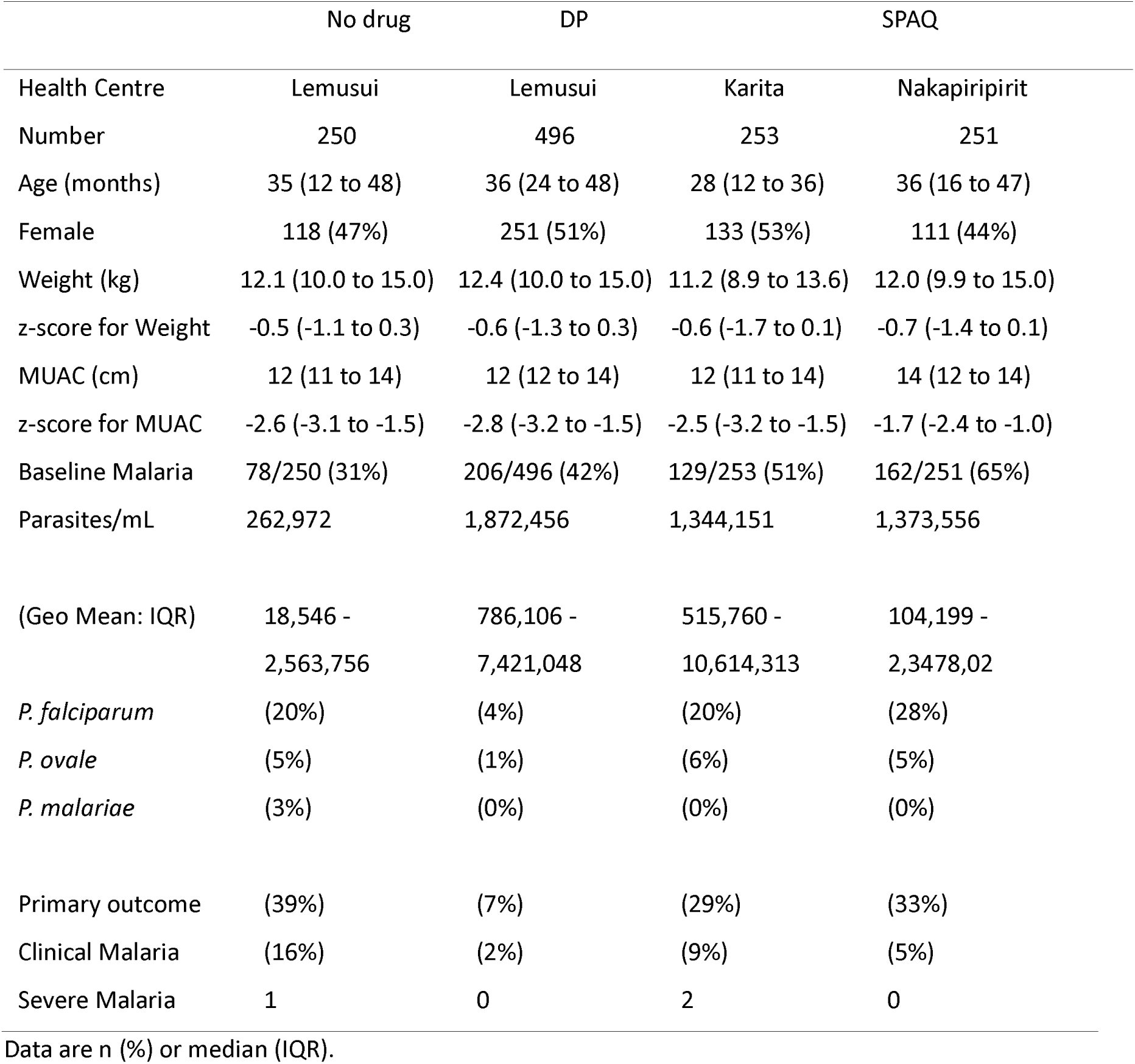
Baseline demographics, malaria prevalence and major outcomes.

### Baseline malaria prevalence

At baseline 561 (45%) children were PCR malaria positive (Figure 1); *P. falciparum* 85% (monoinfections 70%, mixed 15%), *P. ovale* 19% and *P. malariae* in 4.6%. No P. vivax was detected. Mixed *P. ovale* and *P. falciparum* infections were 1.70 (95%CI: 1.29 to 2.24) times more frequent than expected by chance (p=0.0002). The median (IQR) PCR estimated parasite density was 455,870 (12,495 to 6,414,733) genomes/mL.

### Chemopreventive efficacy

The primary outcome of “any malaria infection” occurred in 289 children (Table 1). Unadjusted prevalences on D0 and D28 are shown in Figure 2. After adjustment for baseline malaria positivity, enrolment date, age, and health center, the relative reduction (95% CI) in any malaria infection by day 28 compared to no SMC was 56% (35 to 70%) for SPAQ and 84% (77 to 89%) for DP. For *P. falciparum* chemoprevention efficacies for SPAQ and DP were 46% (11-68%) and 86% (75-92%) respectively. For *P. ovale* and *P. malariae* together, day 28 chemoprevention efficacies were similar, albeit with larger uncertainties: SPAQ 92% (54 to 98%) and DP 94% (80 to 98%). For *P. ovale* specifically (both species) the chemoprevention efficacies were SPAQ 79% (35 to 93%) and DP 96% (84 to 99%). As a sensitivity analysis we re-estimated DP chemoprevention efficacy, removing the children who may have received DP incorrectly from the control group; the estimate was largely unchanged: 87% (95%CI 80 to 91%) chemopreventive efficacy.

**Figure 2:**
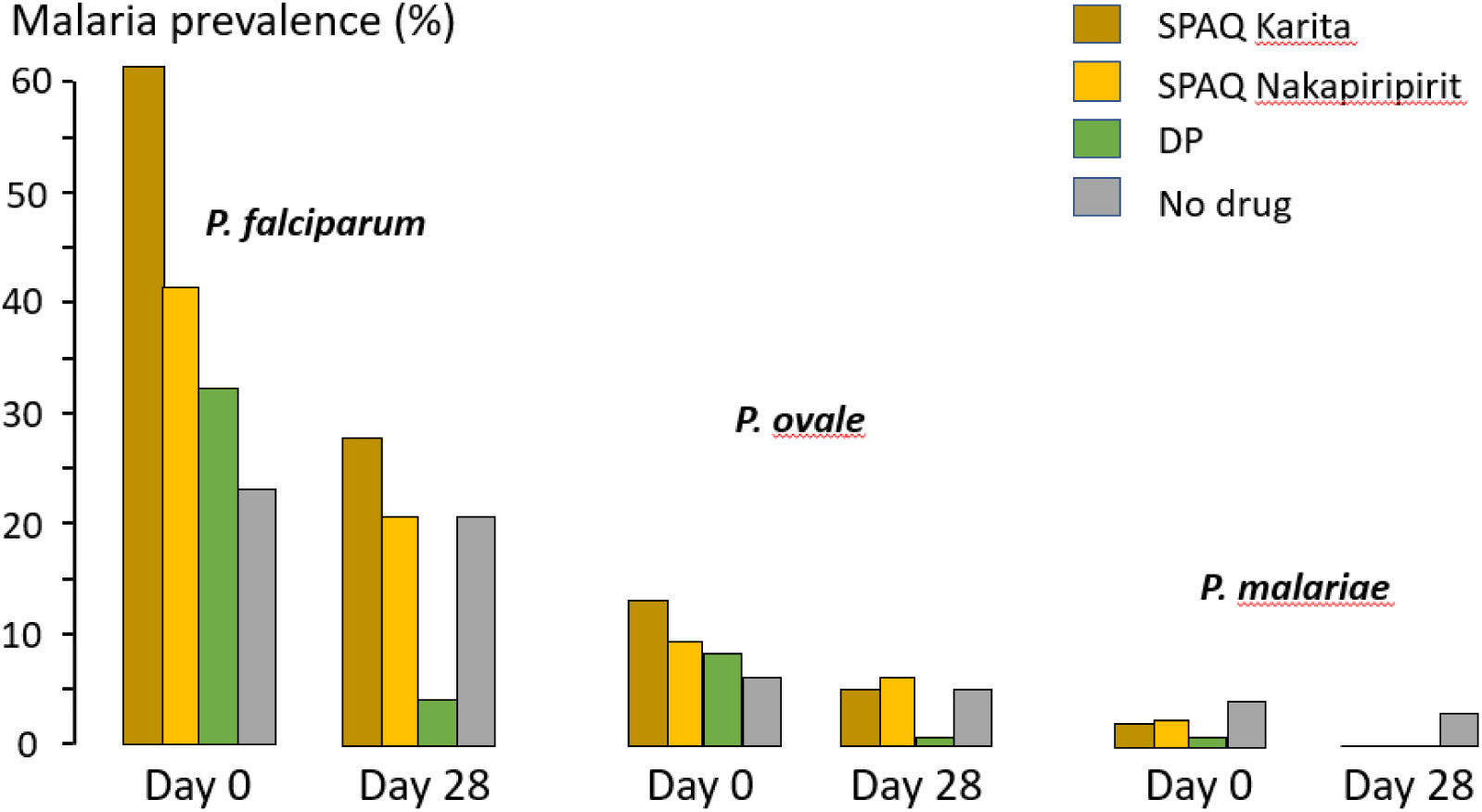
Overall prevalence of malaria on enrolment (D0) and day 28 (D28), based on either slide positivity or qPCR positivity (excluding infections with insufficient DNA for PCR). SPAQ: sulfadoxine-pyrimethamine, DP: dihydroartemisinin-piperaquine. Alt text for Figure 2: This figure shows histograms of the malaria prevalence before (day 0) and 28 days after the chemoprevention was given divided by the three malaria species. The results for SPAQ are divided by the two sites, and the other site is separated by DP and no drug

### Clinical outcomes

Symptomatic malaria developed in 98 children. Five developed clinical malaria within 24 hours. Their D0 geometric mean estimated PCR parasite count was 23,941 genomes/µL (range 1738 to 115,083/µL). As these infections were presumably developing at enrollment, these five children were excluded from the breakthrough infections calculation. There were 13 symptomatic infections in the DP group, 38 in the SPAQ group (2 developed severe malaria) and 42 in the control group (1 developed severe malaria) (Table 1). One child had two separate episodes of severe malaria requiring parenteral artesunate treatment 20 days apart. Under the same multivariable model as for the chemoprevention assessment, the relative reduction in all symptomatic malaria for SPAQ was 60% (27 to 78%) and for DP was 86% (74 to 92%). Children who developed intercurrent symptomatic malaria were 2.8 times (95%CI 1.8 to 4.4) times more likely to have a positive baseline PCR than those who did not. Median (IQR) intervals from D2 to symptomatic malaria presentation were control group 13 (6 to 21) days, DP group 16 (7 to 21) days and SPAQ group 7 (5 to 14) days. The probability of developing clinical malaria decreased whereas the probability of being qPCR positive increased with age. Different temporal patterns were apparent in the different health centers.

### Pharmacokinetics

2686 filter paper DBS were analysed. Because of rapid biotransformation, amodiaquine was barely detectable at day 7 (median 2.0 ng/mL [IQR 0-4.2]). In four children amodiaquine concentrations at D28 were >5ng/mL or higher than at D7, which suggests they had just received amodiaquine. The median (IQR) capillary blood desethylamodiaquine concentrations at D7 and D28 were 157 (117 to 210) ng/mL and 30.8 (18.9 to 48.4) ng/mL respectively (Figure 3). The corresponding median (IQR) D7 and D28 sulfadoxine concentrations were 29800 (22700 to 38700) ng/mL and 2630 (1583 to 5195; N=328) ng/mL. In 158 (33%) children the D28 sulfadoxine level was below LLOQ. The median (IQR) D7 pyrimethamine concentration was 91.7 (70.5 to 121) ng/mL and, as expected, the majority (62%) of children had D28 levels below LLOQ. Estimated median terminal elimination half-lives (t_1/2_β) were piperaquine: 10.4 (IQR 9.1 to 12.4) days, desethylamodiaquine: 8.8 (7.4 to 10.8) days, and sulfadoxine for children with measurable D28 levels: 5.8 (4.9 to 7.2) days. The median elimination half-life of pyrimethamine cannot be calculated, but must be less than 4 days. There was no clear relationship between age and drug exposure for piperaquine but a very strong relationship for desethylamodiaquine; older children had a three-fold lower D28 geometric mean concentration (20ng/ml in children 5 years old, versus 60 ng/ml in 3 month olds).

**Figure 3:**
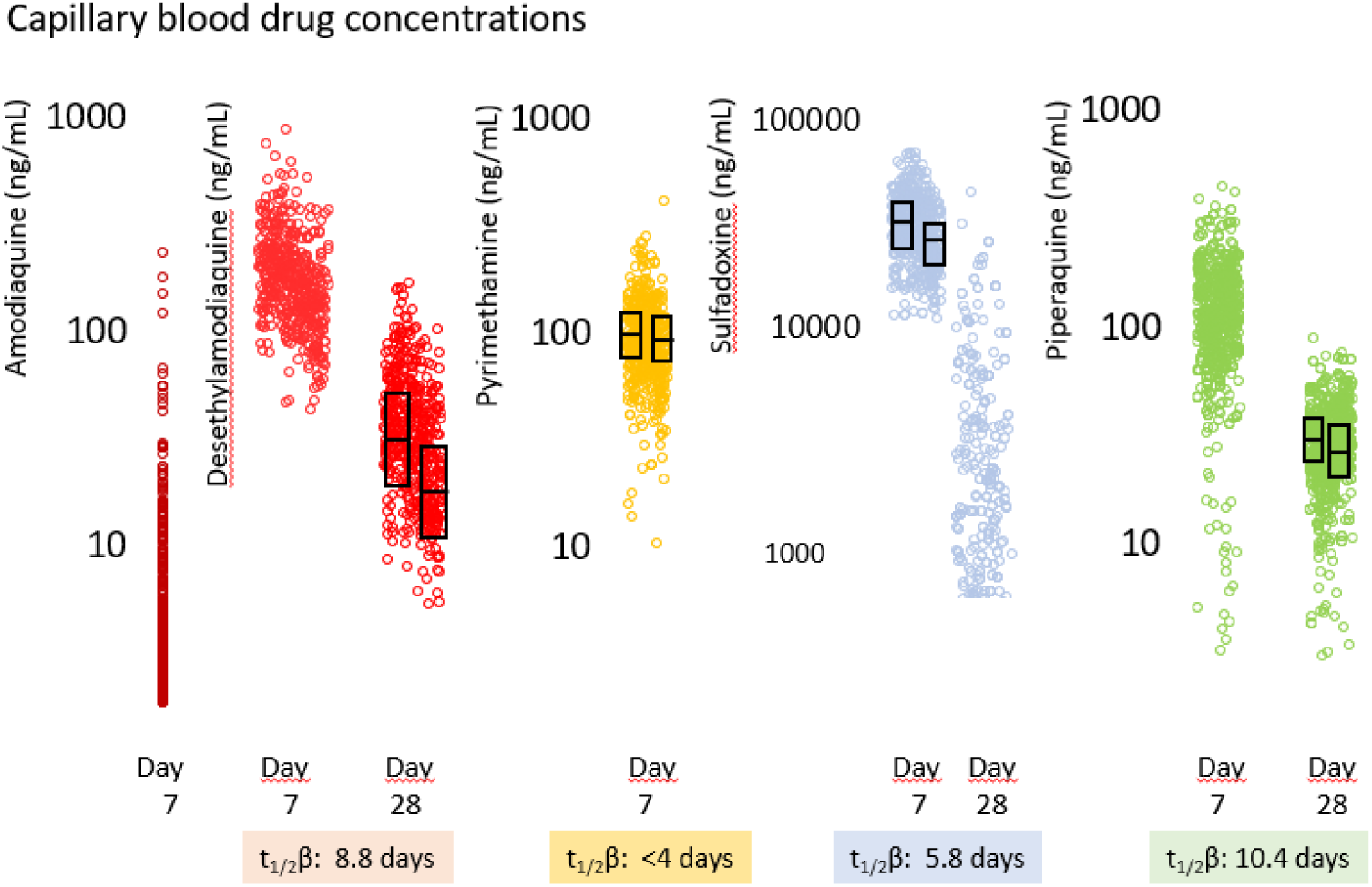
DBS blood concentrations. The boxes show the median and IQR values for children with no malaria parasitemia at D28 on the left and for those with malaria parasitemia detected at D28 on the right for each drug. Estimated terminal elimination half-lives are shown below. Alt text for Figure 3: This plot shows the individual capillary blood concentrations of each drug at day 7 and day 28, the median and IQR values above, and the estimated elimination half-lives below

### Pharmacokinetics and chemopreventive efficacy

Compared with children who did not have breakthrough infections, D28 desethylamodiaquine concentrations were significantly lower in those with breakthrough malaria (22.5 versus 37.0ng/ml, p<0.0001) (Figure 3). Similar differences were observed for the day 7 sulfadoxine and pyrimethamine concentrations, although the levels of the three drugs were correlated. For piperaquine, the differences were less (D28: 26ng/ml in the few breakthroughs versus 31ng/ml). The median D28 desethylamodiaquine concentration (∼30ng/ml) lies on the steep part of the estimated concentration-effect relationship (Figure 4). Although D28 desethylamodiaquine concentrations >50ng/ml prevented malaria parasitemia over one month, most children had much lower concentrations. D7 sulfadoxine, but not D7 pyrimethamine concentrations, were also correlated with chemoprevention efficacy. In a multivariable logistic regression model including all three drugs (D28 desethylamodiaquine, and D7 sulfadoxine and pyrimethamine) and adjusting for baseline parasitemia, age, village, and date of enrolment, the strongest pharmacokinetic predictor of the primary outcome was the D28 desethylamodiaquine concentration (odds ratio of 0.09 for a ten-fold increase, p=0.0001), with a much weaker independent contribution from the D7 sulfadoxine concentration (OR 0.20 for a ten-fold increase, p=0.04) and none from the D7 pyrimethamine concentration.

**Figure 4:**
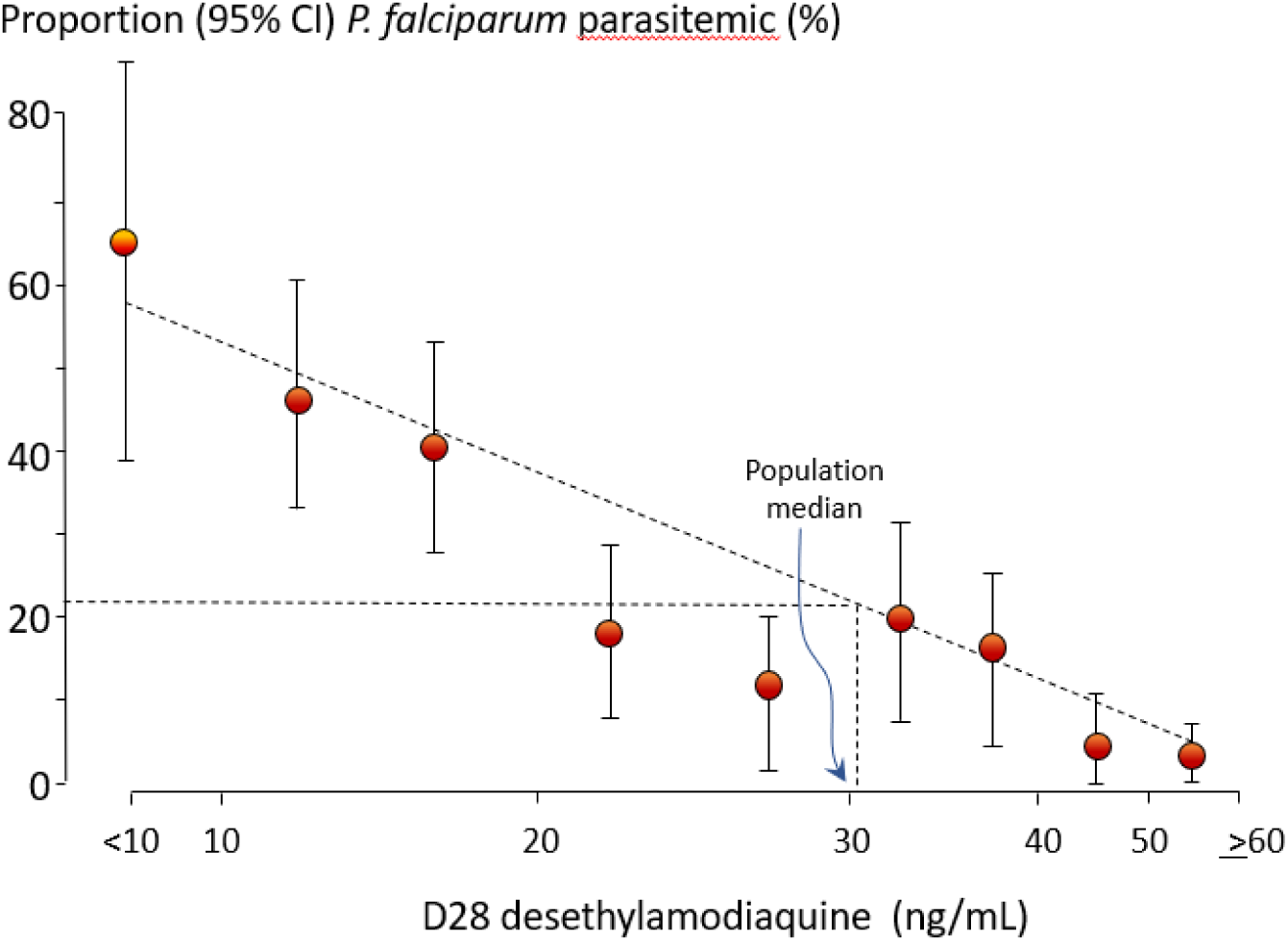
The relationship between D28 parasitemia and D28 desethylamodiaquine concentrations. Median and 95% confidence intervals are shown. Alt text for Figure 4: This shows a straight line fitted graph of the relationship between D28 parasitemia and the D28 desethylamodiaquine concentrations on a log scale (horizontal axis). Median and 95% confidence intervals are shown.

### Molecular markers of antimalarial drug resistance

*P. falciparum* crt mutations at position 76 cause chloroquine resistance [20]. All 260 genotyped *P. falciparum* parasites were “wild type” (haplotype CVMNK) at positions 72-76, and so should have been 4-aminoquinoline sensitive. This suggests that the true prevalence of *Pf* 76T is less than 1.4% (0.975 quantile of a Beta [1,261] distribution). For the *Pfcrt* “downstream” mutations (some implicated in piperaquine resistance) the most frequent haplotype was wild type at positions 93, 97, 145, 218, 343, 353 & 356, and mutant at position 219 (S219N). For Pfmdr1 (also implicated in 4-aminoquinoline resistance) the most frequent haplotype was wild type at positions 86, 784, 945, 1034, 1068, 1042, 1246, and 1314, and mutant at position 184 (Y184F). No parasites had plasmepsin 2 duplications. For *Pfdhfr*, where binding site mutations confer antifolate resistance, the most frequent haplotype was wild type at positions 16 and 164, and mutant at 51(I), 59(R), and 108 (N). For *Pfdhps*, where mutations confer sulfonamide resistance, the most frequent haplotype was wild type at positions 431, 436, 581 and 613, and mutant at 437(G), and 540(E). Thus, for these SP drug target genes, the most frequent haplotype was the “quintuple mutation” haplotype. There was sufficient DNA for *Pf* K13 genotyping of 139 baseline samples; 39 (28%) were “wild type”, 31(22%) were A675V, 25(18%) were C469Y, 37 (27%) were K189T and there were 7 other mutations outside the “propeller” region. Thus 40% of *P. falciparum* parasites had K13 mutations associated with slow parasite clearance after ACT treatment (artemisinin resistance) [21].

## Discussion

The primary objective of SMC deployment is to prevent malaria illness and death by giving at-risk healthy children slowly eliminated antimalarial drugs for several months each year. WHO recommends that SMC should have at least 75% efficacy assessed at 28 days [4], although their definition of preventive efficacy is unclear. Deploying SMC represents a substantial investment of human and financial resources for National Malaria Control Programmes so it is essential that SMC is cost-effective [2]. In this study area in Northern Uganda, where SMC implementation was beginning, 17% of the 250 children in the control group developed symptomatic malaria and one developed severe malaria within 28 days of observation. This reflects the intense transmission and burden of malaria [12]. Assuming no unmeasured confounding, the preventive efficacy of SPAQ with current dosing was weak. SPAQ often failed to clear parasitemia at all, 7.5% of children had breakthrough malaria despite full dosing, two children developed severe malaria, and *P. falciparum* (but not *P. ovale*) often grew through high blood concentrations of desethylamodiaquine, pyrimethamine and sulfadoxine. Overall SPAQ reduced the incidence of symptomatic falciparum malaria in the four week study period by about 60% and reduced the prevalence of malaria parasitemia by just over half compared with the contemporary untreated control group. In comparison DP reduced symptomatic falciparum malaria incidence by 86%, with similar reductions in the prevalence of malaria parasitemia. None of the few DP breakthrough infections developed severe malaria. Overall DP was about four times more effective as malaria chemoprevention compared with SPAQ. DP chemoprevention was similar to SPAQ in preventing recurrent *P. ovale* infections, which comprised about one fifth of the malaria infections overall. But neither drug approached the 100% protective efficacy required of chemoprophylaxis.

Using the same pharmacometric antimalarial resistance monitoring (PARM) approach [13] as in the present study, we showed recently that SPAQ SMC also has modest parasitological efficacy in Northern Mozambique (unpublished observations). In that study area the antimalarial drug susceptibility profile is similar to that in Uganda, although there is no artemisinin resistance reported yet in Mozambique. As in Uganda *P. falciparum* parasites were often able to grow through relatively high SPAQ drug concentrations and often produced mature, microscopy detectable and presumably transmissible, gametocytes. In Uganda the contribution of SP to chemoprevention, based on the exposure-response assessment, was weak -and could not be shown at all in Mozambique. In contrast, although SPAQ had only moderate chemoprevention parasitological efficacy overall, there was a clear exposure-related chemopreventive effect of desethylamodiaquine. The rapid elimination of desethylamodiaquine (the active metabolite of amodiaquine) was an important contributor to the low overall parasitological efficacy of SPAQ [18, 22, 23]. This reflects the relatively young age of these subjects in comparison to previous pharmacokinetic studies. While the exposure-response relationship predicts that doubling of desethylamodiaquine exposures should provide high chemopreventive efficacy (comparable to that of DP), it is unlikely that doubling the amodiaquine dose would be well tolerated, and the alternative of increasing the frequency of drug administration would be operationally challenging, and potentially more toxic [24].

The high level of clinical and parasitological *P. falciparum* SPAQ chemoprevention failure in this study is not explained adequately by the known molecular markers of drug resistance [20]. For SP resistance the main parasite haplotype was the “quintuple” mutant (triple dhfr and double dhps mutations), which should reduce but not abolish entirely the antimalarial effect of the sulfadoxine-pyrimethamine component [20]. This pattern is widespread in East Africa [25]. Nevertheless, amodiaquine alone should still have provided adequate chemoprevention. All genotyped *P. falciparum* parasites were *Pfcrt* “wild type” at position 76 (haplotype CVMNK). Mutation (*Pfcrt* 76T) at this position is the key causal mutation conferring 4-aminoquinoline resistance [20, 26–28]. *Pfcrt* 76K (the “wild type”) should confer susceptibility to chloroquine and amodiaquine. Loss of the key “chloroquine resistance mutation” may have reached fixation in Southeast Africa [28]. Other aminoquinoline resistance mechanisms may be present, or the relatively low desethylamodiaquine concentrations weeks after administration of these doses doses never provide adequate chemosuppression. But it does not explain how some parasites could grow in the presence of high desethylamodiaquine concentrations. In contrast to SPAQ, piperaquine provided well tolerated and relatively effective chemoprevention. The high prevalence of K13 mutations conferring artemisinin resistance in Uganda [14] does not affect DP chemopreventive efficacy against newly acquired infections as the rapidly eliminated dihydroartemisinin does not contribute to the chemoprevention.

Therapeutic efficacy studies traditionally rely on good quality microscopy for parasitological assessment. In East Africa the previously unappreciated high prevalence of *P. ovale* and the difficulty in speciating ring stage parasitemias means that microscopy also has lower specificity than PCR. In this study the majority of recurrent infections were detectable by microscopy, and one third of the D28 *P. falciparum* parasitemias were associated with patent gametocytemia. This suggests that approximately one third of the recurrent falciparum malaria infections were transmissible which reflects the selection pressure on drug resistance. It should be noted that microscopy in this study was performed by expert microscopists who knew the PCR results were positive, so performance is likely to be less good in routine practice or monitoring.

This study has several important limitations and weaknesses. No analytical plan was made before the study. Although there was pseudo-randomisation in one village between DP and no drug, this should be regarded as an observational study. There were significant differences in the baseline prevalence of malaria between the treatment arms, so the forces of infection during the month of observation were also likely to be different. The reinfection risk in the SPAQ group may therefore have been higher than in the DP group and this might have exaggerated the chemopreventive inferiority of SPAQ. Although study conduct was generally good, the drug concentration measurements suggested incorrect treatment allocation in some children and administration of drugs before rather than after taking the blood samples in others. But this is unlikely to have affected the efficacy estimates significantly. The documentation of clinical efficacy was comprehensive and reliable. Overall, these assessments of efficacy conducted under study conditions likely overestimate field effectiveness where SMC coverage is rarely 100% of the target population.

The clinical and parasitological chemopreventive efficacy of SPAQ in this study in Northern Uganda was disappointing, and contrasts with a recent clinical evaluation from the same area [15]. Asymptomatic *P. falciparum* infections were often not cleared by SPAQ and grew through adequate drug concentrations. Breakthrough infections commonly caused illness (which in two cases was severe). One third of recurrent *P. falciparum* parasitemias were accompanied by patent gametocytemia indicating transmissibility and reflecting the drug resistance selection pressure. In contrast DP chemoprevention was relatively effective and well tolerated, as it has been in other chemoprevention uses [29–32]. In addition to recommending widescale deployment of SMC, WHO has also recommended that SP perennial malaria chemoprevention (PMC formerly known as IPTi: i.e. SP treatments given to infants to prevent malaria) should be deployed more widely [33]. This pharmacometrics assessment suggest that PMC with SP only will likely perform very poorly as an antimalarial in East Africa.

The primary objective of chemoprevention is the prevention of illness and death from malaria. It is sometimes argued that antimalarial parasitological efficacy underestimates clinical efficacy in areas of high transmission. Spontaneous resolution of malaria is usual in immune subjects and this overestimates the clinical efficacy of failing antimalarial drugs. Even partial suppression of parasite multiplication may reduce the probability of progressing to severe illness. Twenty years ago this same argument was used to sustain chloroquine well after it had lost efficacy and caused child malaria mortality to rise steeply in Africa. In this study clinical efficacy paralleled parasitological efficacy. Microscopy patent gametocytaemic *P. falciparum* infections commonly followed SPAQ chemoprevention, and parasite densities up to 3% occurred despite adequate desethylamodiaquine concentrations. With such extensive (and expensive) roll-out of untested chemoprevention in areas with high pre-existing levels of drug resistance, it is essential that monitoring of antimalarial chemopreventive efficacy is performed. These results suggest that the utility and cost effectiveness of SPAQ chemoprevention in East Africa should be reviewed [34].

## Competing interests

The authors have no competing interests.

## Data sharing

Pseudonymised participant data used in this analysis are available for access via the WWARN website (https://www.iddo.org/wwarn/accessing-data). Requests for access will be reviewed by a data access committee to ensure that use of data protects the interests of the participants and researchers according to the terms of ethics approval and principles of equitable data sharing. Requests can be submitted by email to via the data access form. WWARN is registered with the Registry of Research Data Repositories. Code used for analysis is available via a github repository: https://github.com/jwatowatson/SMC-Uganda

## Authors’ contributions

CB, AN and JTi conceived of the SMC implementation evaluation with input from KTN, and CR and MRK. AN and RK developed the procedures and wrote the protocol for that study. CE, RK, MO, and DS, coordinated the fieldwork with input from AN, JIN and JO. The pharmacometric study was conceived and designed by NJW, FN and CB. KS and MI conducted the molecular studies, SP conducted the microscopy, and UK and JT conducted the drug analyses. RK, MRK, JN, CE, JAW, CS and NJW had access to and verified the data. JAW and CS conducted the statistical analyses. All authors reviewed the protocol and gave permission for publication.

## Acknowledgements

The authors thank the children and caretakers who participated in the study, the dedicated study staff, village health teams and health facility staff from Amudat and Nakapiripirit districts, and staff at Malaria Consortium and Ministry of Health Uganda. We are also grateful to M. Kiggundu and S. Nsobya for sample collection and processing.

A CC BY or equivalent licence is applied to the author accepted manuscript arising from this submission, in accordance with the grant’s open access conditions.

## Notes

### Competing Interest Statement

The authors have declared no competing interest.

### Clinical Trial

NCT05323721

### Author Declarations

Ethical approval was given by the Mbale Regional Referral Hospital-REC (Ref: MRRH-2022-168) and the Uganda National Council for Science and Technology (UNCST) (Ref: HS2212ES).

