## Supplementary Figures for "Pharmacometric evaluation of amodiaquine-sulfadoxine-pyrimethamine and dihydroartemisinin-piperaquine seasonal malaria chemoprevention in northern Uganda"

**Supplementary Information**

S1: **Catchment area**

All three health facilities in the study area were in the Nakapiripirit district. The catchment area of each health facility was defined as the subcounty or town council where the health facility is located. The selected health facilities were Nakapiripirit Health CenterIII and Karinya Health CenterIII in Nakapiripirit town council where the study drug was SPAQ, and Lemusui Health CenterIII in Moruita sub-county where the study drug was DP. At Lemusui Health Center children were sequentially assigned (2:1) to either DP for the first month or delayed SMC.

**Supplementary figures**

Figure S1:

Dates of enrolment, colored by Health Center.

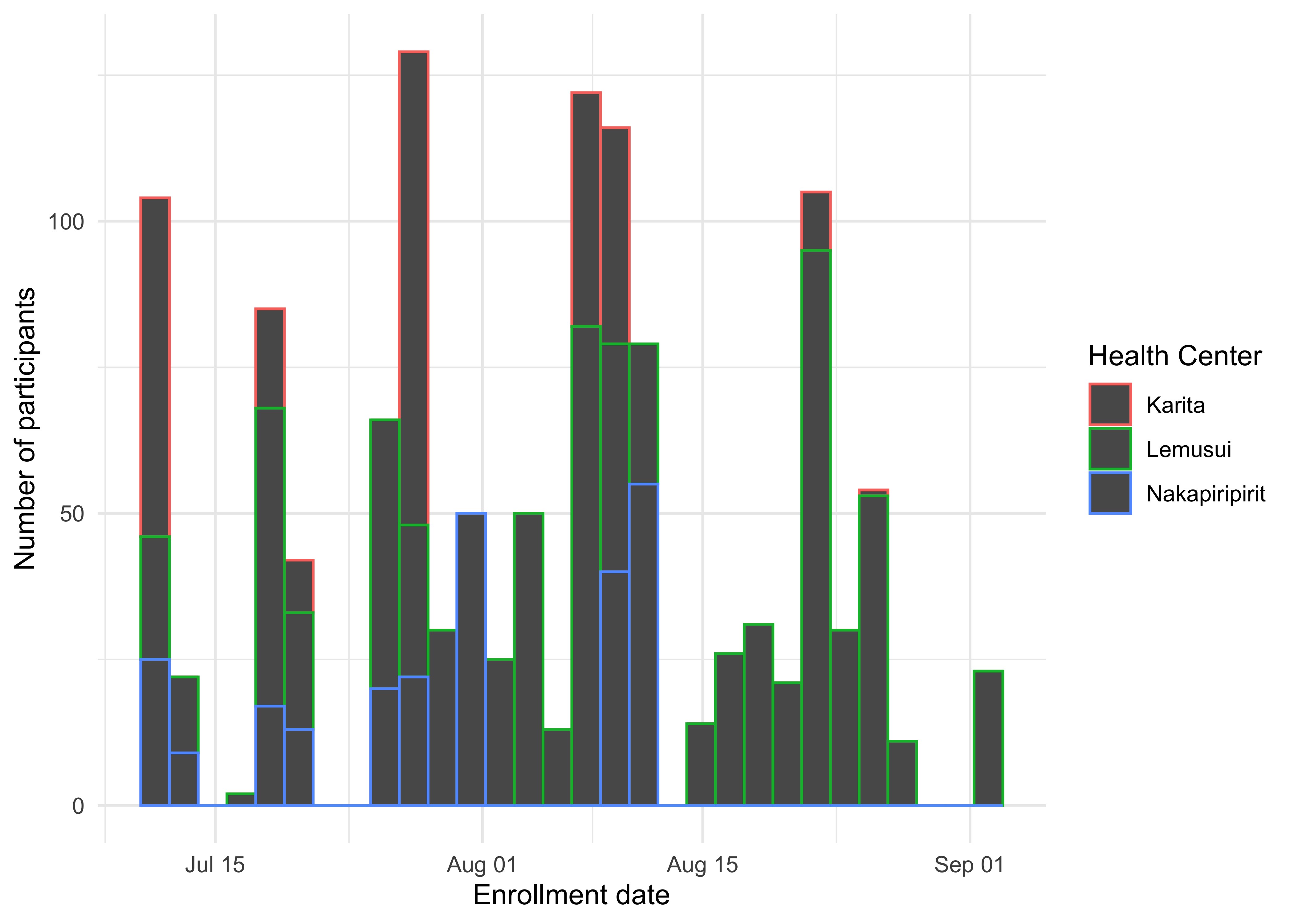

Figure S2:

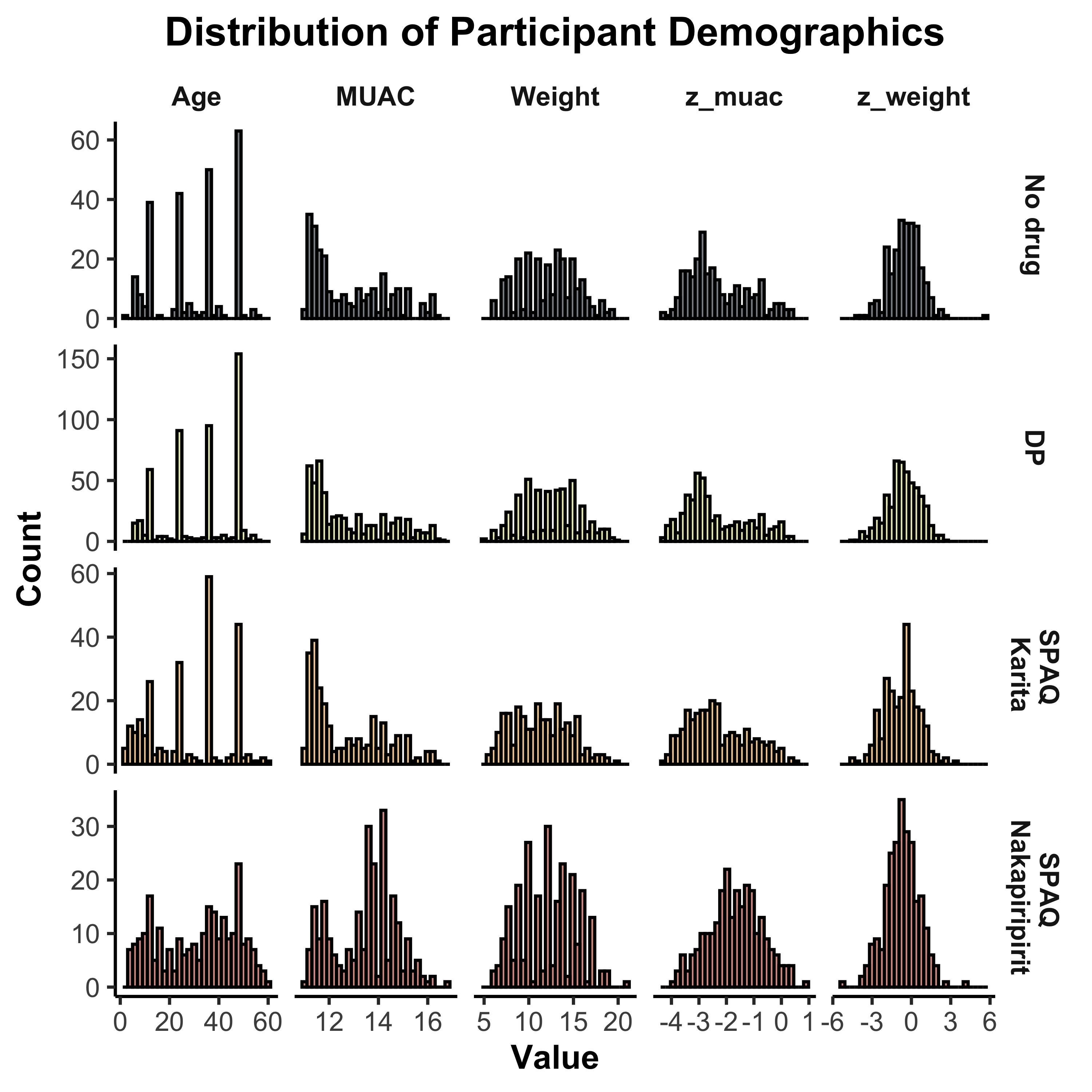

Figure S3:

Parasite densities estimated by qPCR.

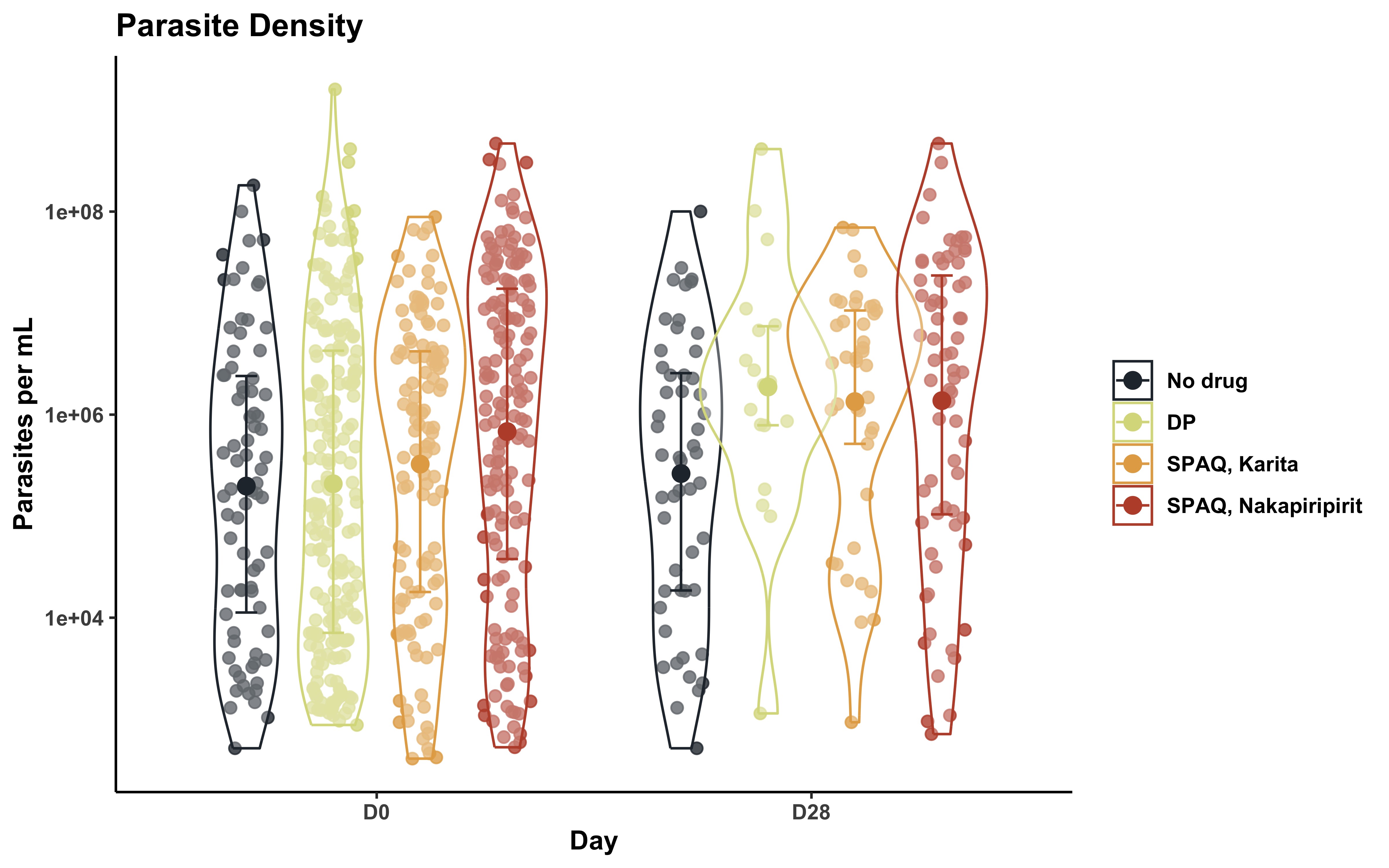

Figure S4:

Relationship between age (months) and the primary outcome of parasitemia between D2 and D28 (left panel) and clinical malaria (right panel). Estimated mean (lines) and 95% confidence intervals (shaded area)

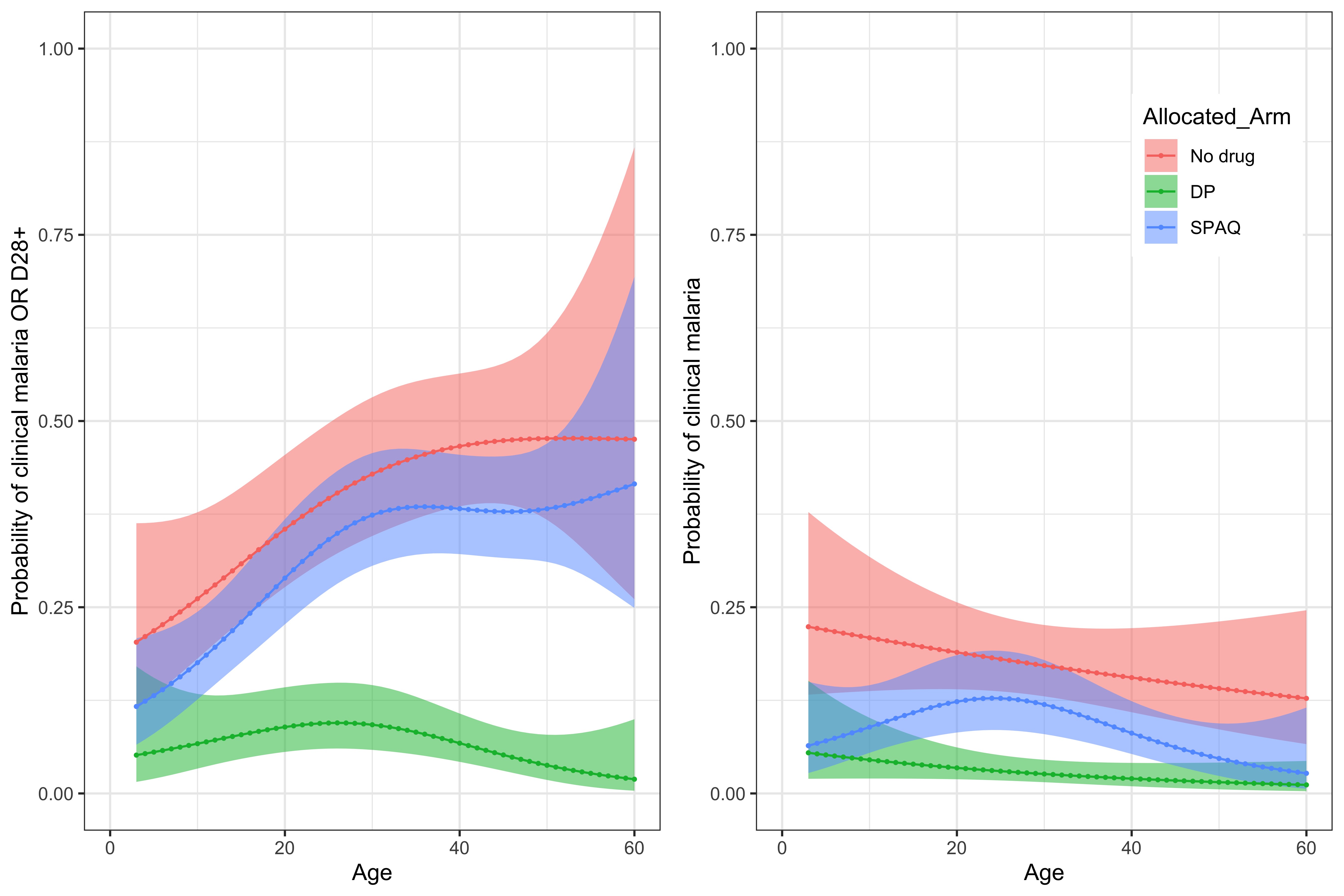

Figure S5:

Relationship between date of enrolment into the study and the occurrence of any malaria parasitemia within 28 days (study primary outcome) (left panel) or clinical malaria (right panel).:
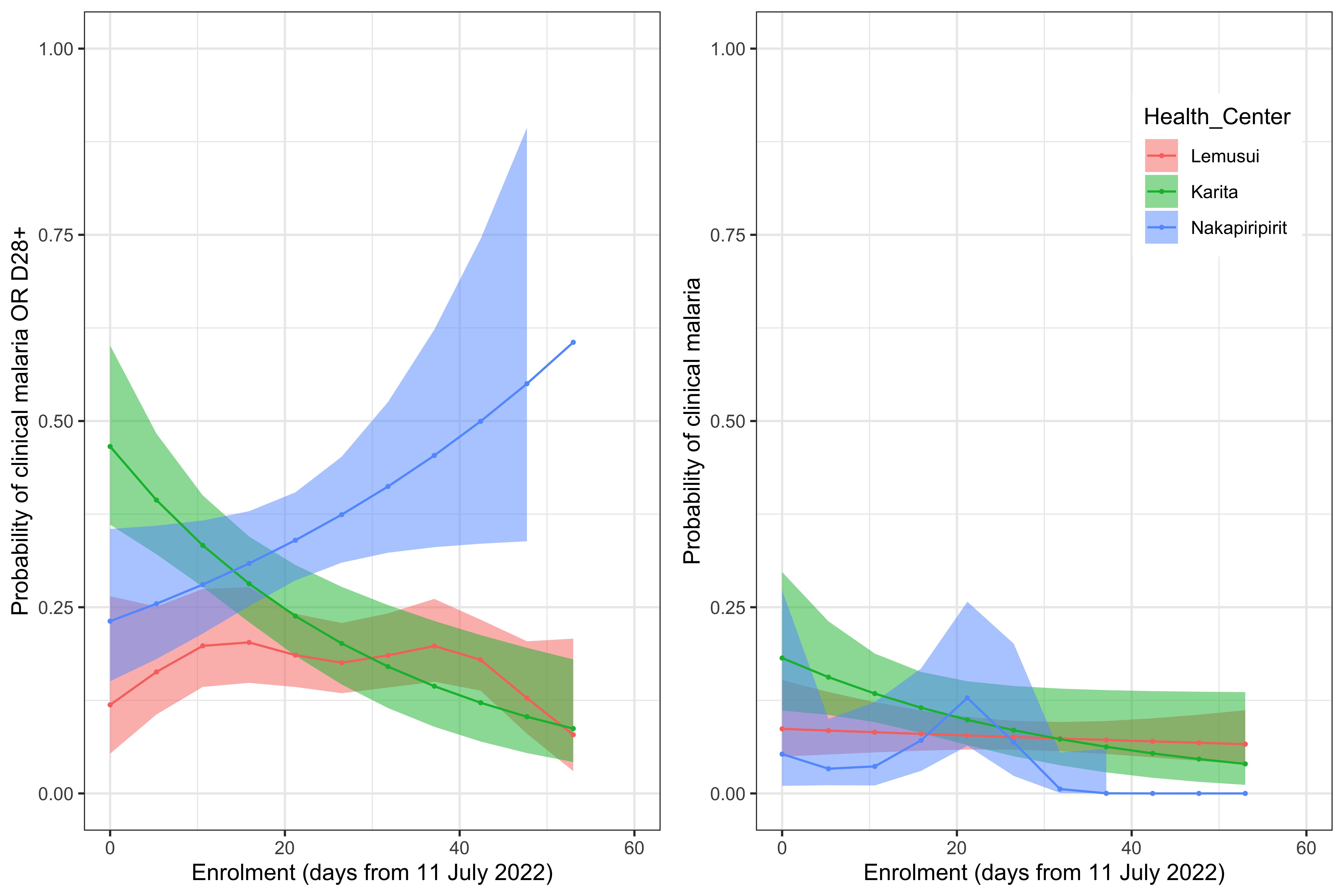

Figure S6:

Relationship between the child’s age (months) and the observed day 28 concentrations of piperaquine (top left) and desethylamodiaquine (top right), sulfadoxine (bottom left) and pyrimethamine (bottom right).

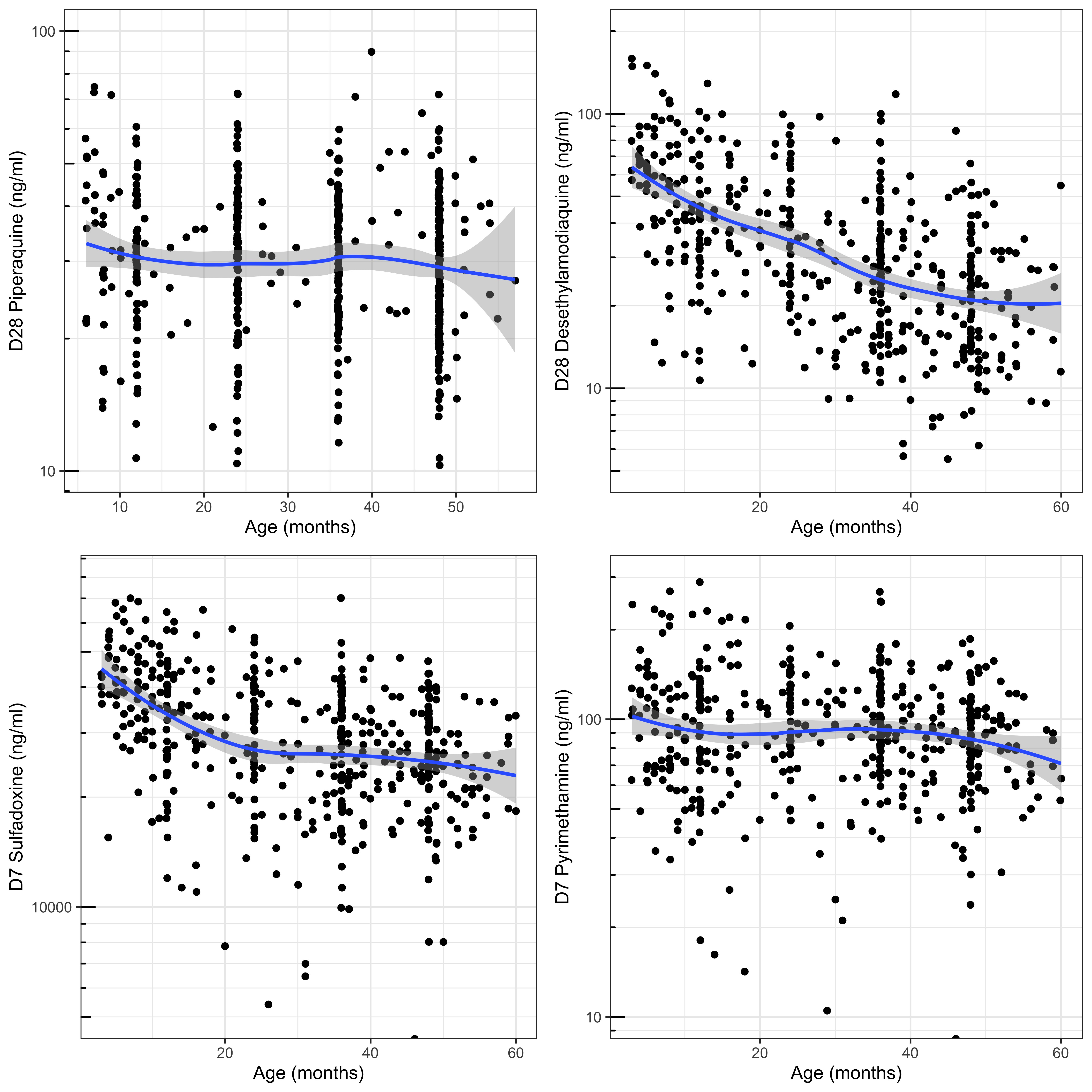

Figure S7: Scatterplots showing correlations between levels of the individual components of SPAQ

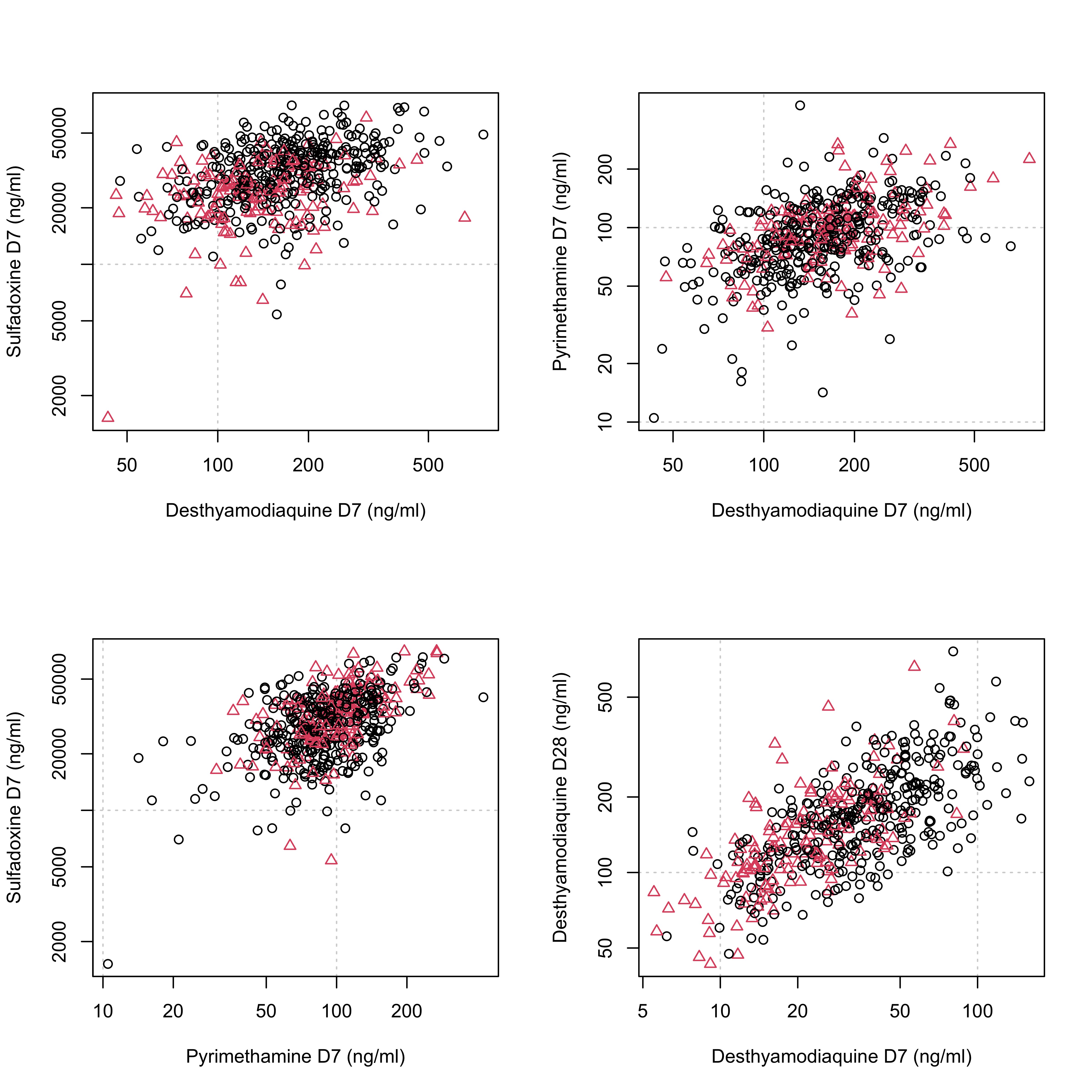
